# Smartphone-based Non-Invasive Classification of Anemia Severity using Deep Learning: A Cross-Sectional Study in Rural India

**DOI:** 10.64898/2026.10.22.26366247

**Authors:** Shrey Desai, Shruti Gadgil, Vineel Abhinav Gottala, Kaushal Jesalpura, Divya Ramnath, Elzbieta Gruzewska, Harshad Purandare, Yun Liu, Umesh Telang, James A Taylor, Rajroshan Sawhney, Richa Tiwari, Dale R. Webster, Shravya Shetty, Avinatan Hassidim, Yossi Matias, Michael Howell, Sethuraman Venkatraman, Bhavna Daryani, Christina Chen, Sujay Kakarmath, Mayank Daswani, Marc Wilson, Raghu Pullakhandam, Swapnil Vispute, Matthew Thompson

## Abstract

Anemia affects over two billion people globally. Screening individuals for anemia in community settings currently involves blood sampling, either a capillary or venous blood sample for running a point of care test or a laboratory grade blood test for hemoglobin, respectively. The need for a blood sample, and laboratory equipment and accessories are barriers to widespread screening for anemia in lower resource settings. We present a non-invasive, artificial intelligence (AI)-driven method for measuring hemoglobin (Hb) and detecting anemia using smartphone-captured images. We collected images of conjunctivae, as well as demographic and clinical data from patients seeking care at SEWA Rural hospital in rural, indigenous population of Gujarat, India. Ground truth Hb values were obtained using venous blood on a Sysmex XN-550 autoanalyser. Age and gender specific WHO reference cutoffs were used for classification of anemia severity. The dataset (n=5,176) was partitioned (70:15:15 ratio) for training (n = 3,623), validation (n = 776) and testing (n = 777). The deep learning based AI model used age, gender, pregnancy status and images of conjunctivae and predicted Hb values which were thresholded for various anemia classification tasks. Overall Hb prediction mean absolute error was 1.12 g/dL.The sensitivity and specificity of the model for classification of any anemia were 70% (95%CI 66-74) and 70% (95%CI 64-76) respectively. Sensitivity (90%, 95% CI 82-94) and specificity (80%, 95% CI 77-83) were higher for detecting severe anemia versus mild, moderate, or no anemia. The accuracy of the model was sustained in a variety of lighting conditions, skin tone and smartphone types. Our findings demonstrate the viability of smartphone based detection of anemia, especially severe anemia, in lower-resource settings and motivate further improvements and validation for use in community settings. The dataset comprising conjunctiva, tongue, nail bed images and smartphone-captured photoplethysmogram (PPG) signal is also released for community research use.

## Introduction

Anemia is a global health problem characterized by insufficient blood hemoglobin (Hb) concentrations to meet physiological needs (1). It disproportionately impacts vulnerable populations: globally, 40% of children under five years of age, 37% of pregnant women, and 30% of all women of reproductive age are affected (2, 3, 4). In India, the burden is particularly high, with the National Family Health Survey (NFHS-5) reporting prevalence rates of anemia as high as 67% in children and 57% in women (5).

Despite its high prevalence, anemia remains underdiagnosed particularly in lower resource settings, due to the lack of awareness, the relative inaccuracy of clinical features, and challenges of accessing hemoglobin tests using automated hematology analysers (6, 7, 8). There is strong evidence for interventions to correct anemia when it is detected, underscoring the value of detection particularly among women and children (9, 10). Although several point of care tests using fingerstick capillary blood samples are in use, these invasive methods require skilled personnel, supply-chain logistics, and reagent stability across diverse environmental conditions, hindering their scaling and sustained implementation in rural healthcare settings (11–15). Non-invasive hemoglobin estimation methods therefore potentially improves the last mile reach of hemoglobin testing, and could be effective tools in anemia control programmes in low and middle income countries (16).

The widespread use of smartphones has prompted several efforts to utilize sensors such as the camera to detect pallor in various areas of the body as a way of estimating Hb (16). However, research in this area has been limited by small, homogenous datasets (often of limited availability to the wider research community) or lack rigorous clinical laboratory reference standards; these have produced encouraging but limited evidence of accuracy (17–24). This study addresses these gaps by first developing a multimodal AI model trained on a large, rural Indian population, and second introducing and making available the associated dataset^25^.

## Methods

### Study Design & Setting

A prospective cross-sectional study was conducted in rural indigenous areas in Gujarat, India by the Society for Education, Welfare and Action–Rural (SEWA Rural), a voluntary development organization involved in health and development activities. The SEWA Rural hospital delivers outpatient and inpatient secondary level health services for adults and children across a range of specialties, and is accessed by individuals from more than 2,500 surrounding villages.

Before initiating the study, expectations about the target product profile, consisting of performance of eventual model and solution design were elicited from multiple stakeholders. These stakeholders included state program officers and public health practitioners and not for profit organizations. These deliberations, informed by other organizations involved in this area, concluded that given the more accessible and non-invasive nature of the target smartphone AI technology, sensitivity and specificity of at least 80% each for various categories of anemia, especially for severe anemia, would meet minimum performance for meaningful use (26). The model also should provide predictions within 30 seconds of inputs, preferably without internet connectivity, and the inputs required for obtaining predictions should be feasible for a community health worker to obtain in community settings after minimal training using a range of low cost smartphones.

### Participants and sampling

Eligible participants were adults, pregnant women and children (1 to 89 years of age) who had a complete blood count (CBC) ordered by a clinician during routine outpatient and inpatient care. Once CBC results were available to the research team, a list of potentially eligible participants were approached and informed about the study in a language of their choice. All participants (and parents/caregivers, where appropriate) provided written informed consent. Potential participants were selected from four categories of Hb level (normal, and mild, moderate, and severe anemia) based on the CBC result, with attempts were made to enrich for patients with severe anemia to improve data diversity for AI model development. Participants who had any material on nailbeds (e.g henna) which could not be removed, or who could not provide consent for the study, were not enrolled.

After the development of the final model, its performance was assessed on both the held-out test set (described below) and a field-representative validation dataset. This field-representative validation dataset included images obtained in settings within SEWA Rural hospital campus that mimic community settings by the same set of research staff using the same aforementioned procedures. The settings within SEWA Rural hospital campus included various indoor and outdoor sites with a variety of lighting, rooms, and wall colors. The participants were selected in the same manner as for the training, validation & testing datasets.

### Procedures

Participant demographics and medical history, including details of health care encounter, pregnancy status and presence of symptoms were collected by research staff based on self-report (or from parent/caregivers) after obtaining an informed consent. A member of research staff measured vital signs including height and weight, temperature (Medtech TMP11 Portable Digital Thermometer, Medtech Life Pvt Ltd, Mumbai, India), heart rate and oxygen saturation (OXYGARD, Pulse Oximeter OG 05, Medtech Life Pvt Ltd, Mumbai, India), and blood pressure (adults only) (Novacheck BP-18, Medtech Life Pvt Ltd, Mumbai, India). Skin tone was measured on the forehead using a spectrophotometer (CAPSURE, Pantone RM200QC Portable Spectrocolorimeter, Panton, Carlstadt, NJ, USA), whereby the L* and b* parameters were converted to an individual typology angle (ITA°) as defined by the function [arctan(L* - 50)/b*] x 180/3.14159 (27). Further, based on the ITA° values, skin tones were categorized as very light (>55°), light (41° to <55°), intermediate (28° to <41°), tan (10° to <28°), brown (−30° to <10°), and dark (<−30°). This standardized classification was used to categorize participants according to their measured skin pigmentation. The research staff also examined the conjunctivae, tongue and nail beds for signs of anemia; subsequently, the staff noted the presence or absence of anemia based on their clinical examination. Data collection occurred in an interior room where ambient lighting was recorded daily (TESTO 540 Pocket sized Luxmeter, Testo, West Chester, USA). Data was recorded using a digital data collection tool to improve data consistency, completeness and quality.

For each patient, one of three smartphones were used for data collection: Samsung A05, Samsung A14 & Samsung S24 (Samsung Electronics Co Ltd, Suwon, South Korea) to provide a range of image quality based on device cost (low to high respectively). Investigators obtained images of conjunctiva from one eye while the participant gently pulled their lower eyelid, tongue in a protruding manner, fingernail views (from any one hand) with the hand pronated flat on a surface (“open hand”) as well as supinated with fingers gently flexed(“closed hand”), and a 30 second video of photoplethysmogram (PPG) from the index finger obtained with finger placed over the camera. A sheet of white paper was used as a reference object and placed above the eyeball while taking an image or underneath the hand for fingernail images (**Appendix 1**). The camera flash was used consistently for all image acquisitions, irrespective of ambient lighting conditions, to provide a more uniform illumination across subjects and minimize variations in image intensity, color, and shadows arising from differences in ambient lighting. In cases where individuals had removable materials (e.g nail polish) on their index finger, these were removed using nail polish remover.

A point of care device was also used to measure Hb (HemoCue Hb 301 System, HemoCue AB, Angelholm, Sweden) using blood remaining from the venous sample provided for their CBC, and using capillary samples obtained using a finger prick (given this is the standard practice in community settings). The reference Hb test was performed at the SEWA Rural Laboratory using a Sysmex XN-L series cell counter (Sysmex XN-550, Sysmex Corporation, Kobe, Japan) which employs an internal quality control system to ensure calibration at three levels of Hb (5.5, 12, and 16.5 g/dL) and coefficient of variation of the assay at < 1%. The presence of hemoglobinopathies was noted from self-report and/or previous reports noted in the laboratory records.

All images obtained by the research staff were made available to a pathologist (co-author KJ) through a web interface, who assessed the quality of images. Images that were of low quality or revealed identity of participants were excluded. All data were stored in a secure India based cloud server storage owned by SEWA Rural. A copy of de-identified data was made available to Google for AI model development.

### Algorithm development

To utilise the correlation between Hb concentrations and the chromatic properties of the target body sites (e.g conjunctiva), all images were subjected to a targeted, heuristic-based preprocessing pipeline. Specifically, a localized crop of the conjunctival region was computationally extracted by leveraging the white paper edge visible in the image as a reference alongside Canny edge detection, effectively isolating the region of interest from extraneous peripheral data. The curated dataset was subsequently partitioned into training, validation, and testing subsets utilizing a 70:15:15 split. The field-representative validation dataset was set aside and was independent from the curated dataset. Prior to model ingestion, these cropped 256×256 pixel images were mapped into the CIELAB (L*a*b*) color space to optimize the representation of visually perceptible color differences.

To mitigate the risk of overfitting inherent to modestly sized datasets, a two-stage ResNet-variant architecture was developed based on the residual learning framework proposed by He et al (28). Each stage comprises a singular convolutional block, the internal architecture of which is detailed in **Appendix 2**. The high-dimensional visual feature maps extracted by this ResNet backbone are subsequently concatenated with discrete demographic and clinical metadata - namely, the patient’s age, sex, and pregnancy status. The resulting multimodal feature vector was then fed into a multilayer perceptron (MLP)-based fusion network to produce a continuous-valued estimate of Hb concentration. The complete architectural schematic is provided in **Appendix 3**.

The model performed a continuous regression task to predict absolute Hb levels. The model was trained from a random initialization, without any pretrained weights, and optimized using the Adam optimizer with a batch size of 64, an initial learning rate of 0.02 and an exponential decay schedule with a rate of 0.97. At inference, the model’s Hb predictions were subsequently used for classification of anemia and its grades using age and gender specific hemoglobin thresholds recommended by WHO (29). (**Appendix 4**). The overall model configuration is described in **Appendix 5**.

We anticipated that the dataset would be skewed towards mild-to-moderate anemia cases, even with the attempts made to recruit across all grades of anemia. Using a standard Mean Squared Error (MSE) loss would cause a model to favor this majority group over less common (but clinically important) severe anemia cases. To handle this data imbalance, we applied a two-step optimization strategy. First, a Huber loss function (30) was utilized to balance gradient stability with outlier sensitivity, preventing extreme deviations from disproportionately distorting the model. Second, an inverse bin frequency re-weighting strategy was employed. The continuous range of Hb values was divided into a fixed number of bins, with bin widths determined by the sample distribution. Bins representing the severe range (Hb 1 to 7 g/dL) capture a much smaller fraction of the dataset due to the inherent rarity of these cases. By mathematically scaling the error penalty inversely to the sample density within each bin, the training became cost-sensitive. This method has been investigated as a cost-sensitive strategy for addressing target-distribution imbalance in deep regression (31), and in our setup forces the model to heavily weigh and accurately predict the more severe (and under-represented) samples.

To address systematic overprediction in the regression outputs, a constant bias-correction term was applied to the predicted hemoglobin values, with its magnitude selected based on validation set performance. The corrected predictions were then classified using the WHO-defined hemoglobin thresholds, improving the sensitivity–specificity trade-off across clinically relevant anemia classification tasks.

The modeling pipeline was designed for low-latency inference, ensuring it could eventually run on mid-range smartphones used by community health workers in India. Ablation experiments evaluated the impact of incorporating various additional (non-conjunctiva) image modalities (tongue, nails) and clinical metadata (including symptoms, past medical history) against “baseline” models which included only conjunctiva, age, sex, pregnancy status.

Additional ablation experiments evaluated the impact of incorporating various combinations of image modalities (ie conjunctivae, tongue, open hand, closed hand), PPG data, clinical metadata (e.g age, pregnancy status, gender) and symptoms. Symptoms for adults and children were only included in age specific models (ie adult symptoms were only applied to a model involving the adult subgroup and a binary indicator variable for child versus adult status). The outcomes of these ablation studies were used to select the final model for testing in the test set and field validation set, and to describe the tradeoffs between model input complexity and both accuracy of Hb measurement (ie MAE) and classification of anemia.

### Sample size and statistical analysis

Sample size calculated was limited by lack of current estimates of MAE or SD from existing image recognition or PPG software. Number of participants was at least 2,322 to have a power of 0.9 to detect a mean error (device - ground truth) in Hb measurement that is significantly < 1.0 g/dL, with multiple postulated mean errors and standard deviations of the error (one-sided P value < 0.05). We aimed to enroll a sample to include approximately 25% of participants with no anemia, 25% mild, 25% moderate, and 25% with severe anemia.

Characteristics of the study population, including demographics, occupation, clinical settings were described using descriptive statistics. The performance of algorithms were examined within each subset of normal Hb, and anemia (mild, moderate, severe) using metrics such as R^2^, intra class coefficient (ICC) and Bland Altman plots (32). We also measured mean absolute error (MAE) and its standard deviation (SD), as well as 95% confidence interval (95%CI) of estimated Hb. The 95% confidence interval was computed by bootstrapping, resampling 1,000 times with replacement. A one sample t-test (or Wilcoxon signed rank test if data were skewed) was used to determine if the MAE is less than 1.0 g/dL (one-sided P < 0.025).

Sensitivity and specificity (and 95% CIs) were calculated for *a priori* subgroups, defined by age, sex and pregnancy status using established thresholds for defining mild, moderate and severe anemia. Accuracy of the model was also compared with presence of anemia noted on clinical features, and point of care Hb.

### Failure analysis

We conducted a qualitative error analysis to characterize failure modes beyond aggregate metrics. Particular attention was given to clinically significant errors, especially cases of severe anemia misclassified as non-severe and normal cases incorrectly classified as moderate or severe. Among the 106 ground-truth severe cases in the test set, the model achieved 90% sensitivity for severe versus non-severe classification, corresponding to approximately 11 severe cases being misclassified as non-severe. The misclassified severe cases had ground-truth hemoglobin concentrations of 6.5–7.0 g/dL, close to the severe anemia threshold (7.0 g/dL), and involved pregnant women participants aged > 12 years whose conjunctival images exhibited patchy red regions against a white background. These borderline hemoglobin values, potentially overlapping image features, and limited training examples near the decision boundary may have hindered the model’s ability to distinguish severe from moderate anemia. We also examined adjacent-class errors, including normal–mild, mild–moderate, and moderate–severe misclassifications, which may occur when the predicted Hb lies close to the predefined class thresholds. Representative failure cases were further assessed for image-related factors such as poor illumination, image quality and variations in conjunctival appearance. Although nail and tongue modalities were evaluated, their inclusion did not improve performance, suggesting limited complementary information in the current dataset.

### Ethical considerations

The study received approval from SEWA Rural’s Institutional Ethics Committee on 31 March 2023 and has been registered with the Clinical Trial Registry of India (CTRI/2023/07/055313). Clearance to conduct the study was also granted by the Health Ministry Screening Committee (HMSC). Written informed consent was received from all the participants.

## Results

### Description of participants

A total of 5,455 adults and children were recruited between July 2024 and November 2025, with a further field-representative validation cohort (n=516) collected between December 2025 and January 2026. Of the initial 5,455 captures, 279 were excluded due to incomplete metadata, images, and/or errors in image crops. Hence, a total of 5,176 captures were used for training (3,623), tuning (776) and testing (777) (**Figure 1**). Participants were distributed across normal Hb (1398, 27.0%), mild anemia (1303, 25.2%), moderate anemia (1764, 34.1%) and severe anemia (711, 13.7%) categories (**Table 1**). The majority of participants (4334/5176, 83.7%) were female, of whom just less than half, 48.1% were currently pregnant, and almost all were recruited from outpatient settings, mainly from obstetrics and gynecology (56.8%) and general medicine (31.5%) clinics.

**Figure 1:**
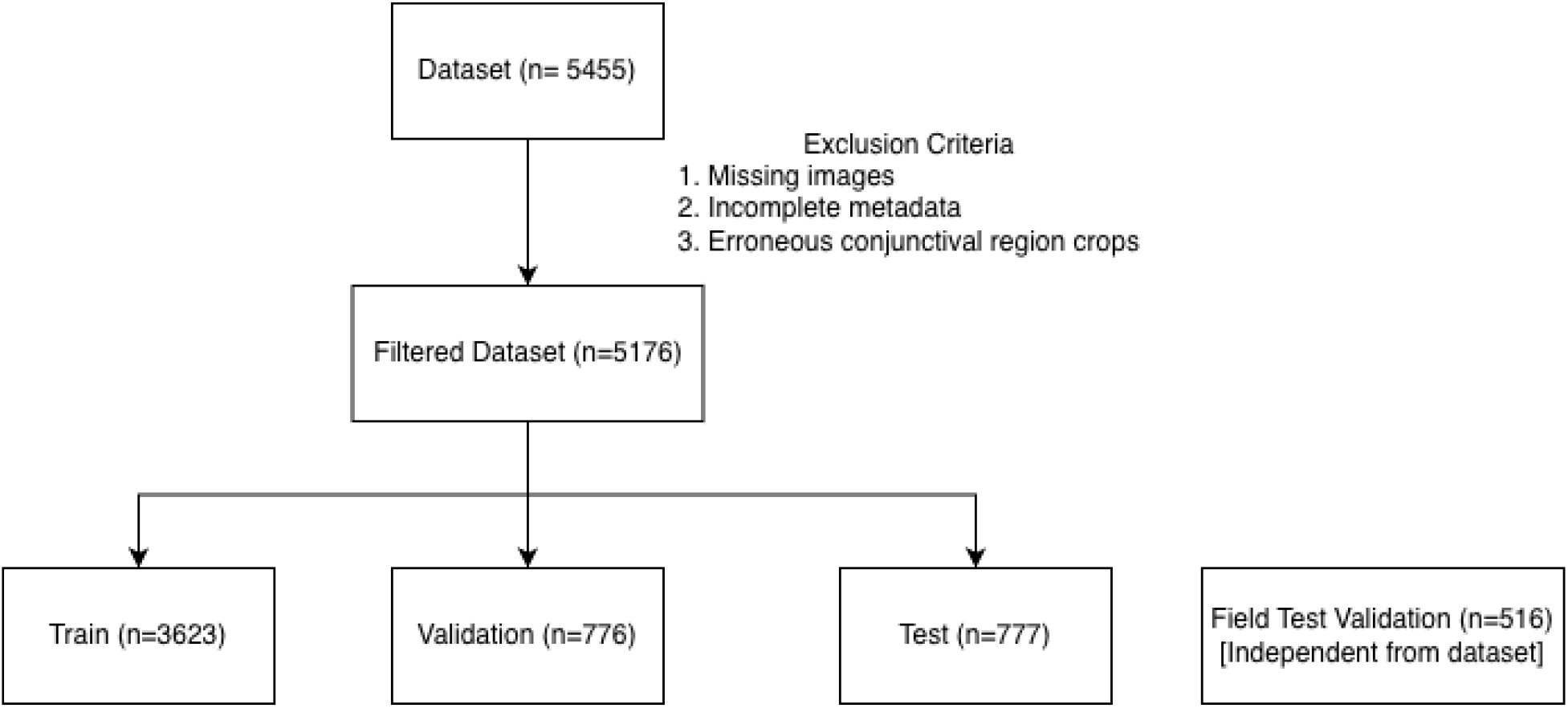
Overview of dataset and splitting

**Table 1:** Characteristics of included population.

|  |  | **Total**<br>**N= 5176**<br>**(100%)** | **Normal Hb**<br>**n=1398**<br>**(27.0%)** | **Mild anemia**<br>**n=1303**<br>**(25.2%)** | **Moderate**<br>**anemia**<br>**n=1764**<br>**(34.1%)** | **Severe**<br>**anemia**<br>**n =711**<br>**(13.7%)** |
| --- | --- | --- | --- | --- | --- | --- |
| **Demographics** |  |  |  |  |  |  |
| **Age (years)** | 0-12 | 531(10.2%) | 90(16.9%) | 89(16.7%) | 318(59.8%) | 34(6.4%) |
|  | 0-19 | 1592(30.7%) | 283(17.7%) | 422(26.5%) | 748(46.9%) | 139(8.7%) |
|  | >19 | 3584(69.2%) | 1115(31.1%) | 1016(24.5%) | 881(28.3%) | 572(15.9%) |
| **Sex** | Female | 4334 (83.7%) | 1162 (26.8%) | 1083 (24.9%) | 1484 (34.2%) | 605 (13.9%) |
|  | Male | 838 (16.2%) | 236 (28.2%) | 218 (26.0%) | 278 (33.2%) | 106 (12.6%) |
|  | Other | 4 (0.1%) | 0 (0.0%) | 2 (50.0%) | 2 (50.0%) | 0 (0.0%) |
| **Details of current health care visit** |  |  |  |  |  |  |
| **Setting** | Outpatient | 5129 (99.1%) | 1392 (27.1%) | 1302 (25.4%) | 1753 (34.2%) | 682 (13.3%) |
|  | Inpatient | 47 (0.9%) | 6 (12.8%) | 1 (2.1%) | 11 (23.4%) | 29 (61.7%) |
| **Service**<br>**attended** | Pediatrics | 253 (4.9%) | 69 (27.3%) | 40 (15.8%) | 113 (44.7%) | 31 (12.3%) |
|  | OBGYN/Antenatal | 2939 (56.8%) | 959 (32.6%) | 797 (27.1%) | 881 (30.0%) | 302 (10.3%) |
|  | Care | 224 (4.3%) | 29 (12.9%) | 35 (12.9%) | 53 (23.7%) | 107 (47.8%) |
|  | Internal Medicine | 1630 (31.5%) | 292 (17.9%) | 401 (24.6%) | 685 (42.0%) | 252 (15.5%) |
|  | General Medicine | 88 (1.7%) | 34 (38.6%) | 25 (28.4%) | 20 (22.7%) | 9 (10.2%) |
|  | General surgery | 39 (0.8%) | 16 (41.0%) | 4 (10.3%) | 8 (20.5%) | 11 (28.2%) |
|  | Ophthalmology | 17 (0.3%) | 4 (23.5%) | 2 (11.8%) | 7 (41.2%) | 3 (17.6%) |
|  | Other |  |  |  |  |  |
| **Medical history** |  |  |  |  |  |  |
| **Currently**<br>**pregnant**<br>*(applies only to*<br>*female*<br>*participants)* | Yes | 2490 (48.1%) | 911 (36.6%) | 724 (29.1%) | 711 (28.6%) | 144 (5.8%) |
|  | No | 1844 (35.6%) | 251 (13.6%) | 359 (19.5%) | 773 (41.9%) | 461 (25.0%) |
| **Smoking**<br>**history** | Current or previous | 57 (1.1%) | 17 (29.8%) | 13 (22.8%) | 8 (14.0%) | 19 (33.3%) |
|  | Never | 5119 (98.9%) | 1381 (27.0%) | 1290 (25.2%) | 1756 (34.3%) | 692 (13.5%) |
| **Alcohol intake\*** | Current or previous | 13 (0.3%) | 0 (0.0%) | 2 (15.4%) | 0 (0.0%) | 11 (84.6%) |
|  | Never | 4264 (82.4%) | 1175 (27.6%) | 1095 (25.7%) | 1453 (34.1%) | 541 (12.7%) |
| **Hypertension\***<br>**(adult)** | Yes | 6 (0.1%) | 2 (33.3%) | 1 (16.7%) | 3 (50.0%) | 0 (0.0%) |
|  | No | 3987 (77.0%) | 1195 (30.0%) | 1000 (25.1%) | 1174 (29.4%) | 618 (15.5%) |
| **Diabetes\***<br>**Mellitus (adult)** | Yes | 20 (0.4%) | 8 (40.0%) | 4 (20.0%) | 4 (20.0%) | 4 (20.0%) |
|  | No | 3973 (76.8%) | 1189 (29.9%) | 997 (25.1%) | 1173 (29.5%) | 614 (15.5%) |
| **Sickle Cell** | Trait | 304 (5.9%) | 83 (27.3%) | 80 (26.3%) | 103 (33.9%) | 38 (12.5%) |
|  | Disease | 655 (12.7%) | 19 (2.9%) | 86 (13.1%) | 442 (67.5%) | 108 (16.5%) |
|  | Negative | 810 (15.7%) | 208 (25.7%) | 202 (24.9%) | 282 (34.8%) | 118 (14.6%) |
|  | Not Known | 3407 (65.8%) | 1078 (31.6%) | 947 (27.8%) | 935 (27.4%) | 447 (13.1%) |
| **Symptoms** |  |  |  |  |  |  |
| **Adults** | Tired or lack of energy | 1352 (26.1%) | 327 (24.2%) | 268 (19.8%) | 356 (26.3%) | 401 (29.7%) |
|  | Shortness of breath at rest or on exertion | 268 (5.2%) | 72 (26.9%) | 42 (15.7%) | 56 (20.9%) | 98 (36.6%) |
|  | Fast heart beat, particularly during exercise | 18 (0.3%) | 3 (16.7%) | 0 (0.0%) | 5 (27.8%) | 10 (55.6%) |
|  | Losing Weight | 58 (1.1%) | 12 (20.7%) | 13 (22.4%) | 19 (32.8%) | 14 (24.1%) |
|  | Chest Pain | 156 (3.0%) | 31 (19.9%) | 27 (17.3%) | 42 (26.9%) | 56 (35.9%) |
|  | Lightheaded or Dizzy | 787 (15.2%) | 168 (21.3%) | 141 (17.9%) | 200 (25.4%) | 278 (35.3%) |
|  | Headache | 891 (17.2%) | 217 (24.4%) | 169 (19.0%) | 236 (26.5%) | 269 (30.2%) |
|  | Leg cramps | 687 (13.3%) | 143 (20.8%) | 128 (18.6%) | 178 (25.9%) | 238 (34.6%) |
|  | Brittle hair or nails | 11 (0.2%) | 0 (0.0%) | 1 (9.1%) | 2 (18.2%) | 8 (72.7%) |
| **Children** | Looks pale | 263 (5.08%) | 17 (6.5%) | 44 (16.7%) | 146 (55.5%) | 56 (21.3%) |
|  | Losing weight/ not gaining weight | 5 (0.09%) | 2 (40.0%) | 0 (0.0%) | 1 (20.0%) | 2 (40.0%) |
|  | More tired or sleepy than usual | 116 (2.24%) | 7 (6.0%) | 27 (23.3%) | 57 (49.1%) | 25 (21.6%) |
|  | Out of breath or difficulty breathing | 16 (0.31%) | 3 (18.8%) | 4 (25.0%) | 7 (43.8%) | 2 (12.5%) |
| **Skin Tone** |  |  |  |  |  |  |
|  | Very light/ light/intermediate | 22 (0.4%) | 6 (27.3%) | 3 (13.6%) | 12 (54.5%) | 1 (4.5%)28 |
|  | Tan | 231 (4.5%) | 57 (24.7%) | 59 (25.5%) | 87 (37.7%) | (12.1%)534 |
|  | Brown | 3946 (76.2%) | 1048 (26.6%) | 1008 (25.5%) | 1356 (34.4%) | (13.5%)148 |
|  | Dark | 977 (18.9%) | 277 (28.4%) | 245 (25.1%) | 307 (31.4%) | (15.1%) |
| **Physical Examination findings** |  |  |  |  |  |  |
| **Nails** | Koilonychia | 114 (2.2%) | 6 (5.3%) | 7 (6.1%) | 30 (26.3%) | 71 (62.3%) |
|  | Clubbing of nails | 180 (3.5%) | 33 (18.3%) | 29 (16.1%) | 58 (32.2%) | 60 (33.3%) |
|  | Brittle or spoon-shape nails | 125 (2.4%) | 3 (2.4%) | 6 (4.8%) | 24 (19.2%) | 92 (73.6%) |
| **Conjunctiva** | Pale conjunctiva | 3334 (64.4%) | 612 (18.4%) | 720 (21.6%) | 1311 (39.3%) | 691 (20.7%) |
| **Tongue** | Pale tongue | 851 (4.8%) | 137 (16.1%) | 129 (15.2%) | 272 (32.0%) | 313 (36.8%) |

*\*The sum of participants across categories may not equal the total sample size due to missing data*

### Estimation of hemoglobin values using AI model and Hemocue point of care test

The proposed model’s prediction of Hb value in the test set (n=777), demonstrated an overall MAE of 1.12 g/dL (**Table 2**). The MAE was lowest in the mild anemia group (0.74 g/dL) and higher in the normal, moderate, and severe groups (1.19 g/dL, 1.23 g/dL, and 1.43 g/dL, respectively). The proportion of predictions within +/-1.0 g/dL was 54%. By category, accuracy was highest in the mild anemia group (71%), followed by the normal group (51%), and lowest in both the moderate and severe anemia groups (47%). **Figure 2** illustrates the Bland-Altman plot evaluating the agreement between ground truth and predicted Hb values. The analysis reveals a mean bias of −0.02 g/dL, demonstrating that the model possesses negligible systematic error. The 95% limits of agreement (LoA) ranged from a lower limit of −2.90 g/dL (−1.96 SD) to an upper limit of +2.85 g/dL (+1.96 SD). The vast majority of the data points fell within these boundaries, demonstrating acceptable statistical consistency across the clinical range of Hb. The MAE for Hemocue point of care device using either venous or capillary samples were broadly similar for overall performance(0.48 g/dL and 0.52 g/dL, respectively) as well as across anemia categories. (**Table 2**).

**Figure 2:**
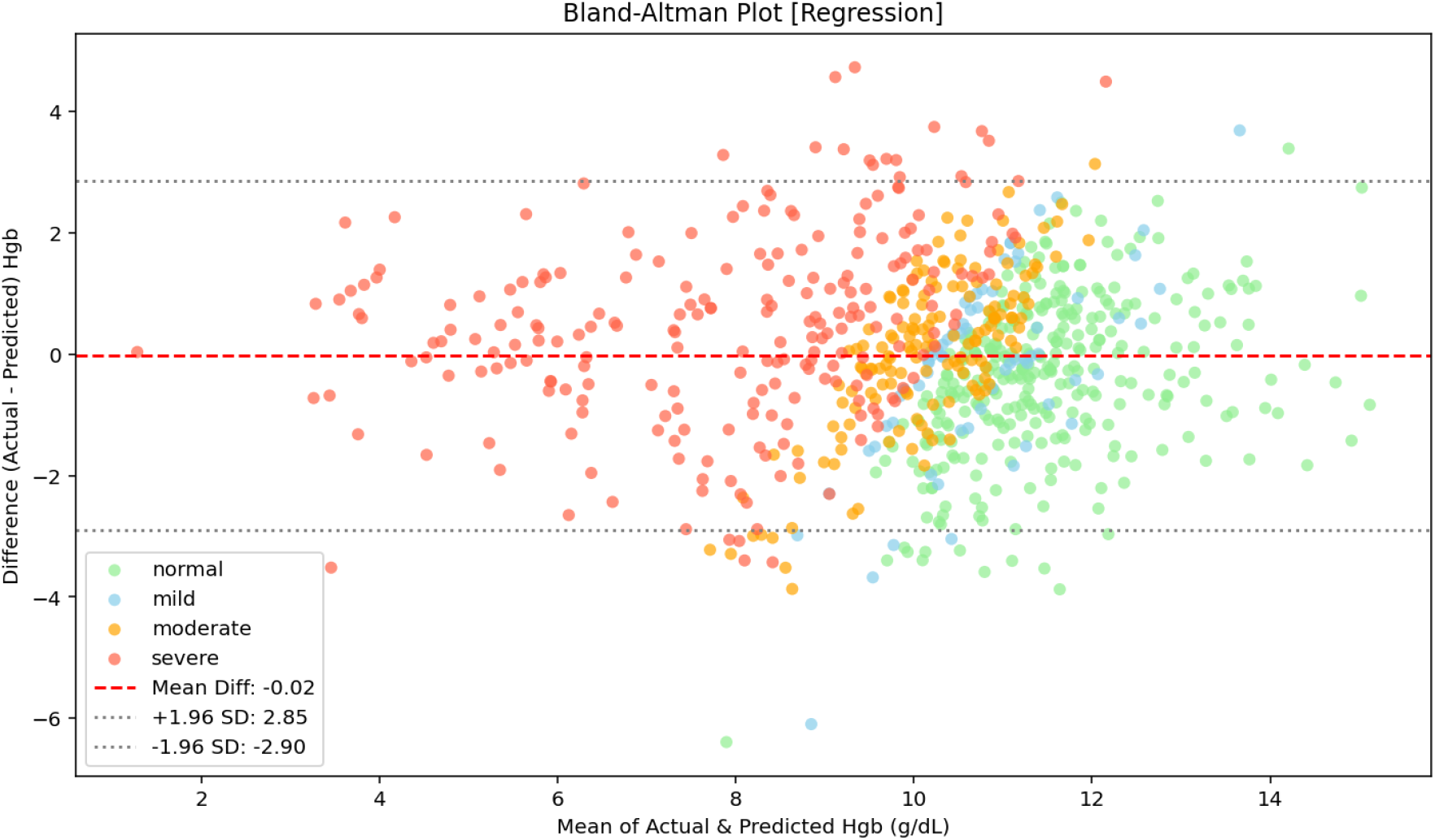
Bland Altman plot of agreement between ResNet predictions of Hb and laboratory ground truth in the test dataset

**Table 2:** Mean Absolute Error (MAE) of Hb estimation by the AI model, as well as Hemocue venous and capillary samples, for the overall population and by severity of anemia in the test dataset (n=777)

|  | AI Model<br>(n=777) |  | Hemocue, venous sample<br>(n=777) |  | Hemocue, capillary sample<br>(n=516) |  |
| --- | --- | --- | --- | --- | --- | --- |
|  | Mean<br>absolute<br>error<br>(g/dL) | Performance<br>accuracy<br>within +/-<br>1.0g/dL Hb | Mean<br>absolute<br>error<br>(g/dL) | Performance<br>accuracy<br>within +/-<br>1.0g/dL Hb | Mean<br>absolute<br>error<br>(g/dL) | Performance<br>accuracy<br>within +/- 1.0g/dL<br>Hb |
| **Overall** | 1.12<br>(n=777) | 54% | 0.48<br>(n=777) | 97% | 0.52<br>(n=516) | 90% |
| **Normal** | 1.19<br>(n=210) | 51% | 0.40<br>(n=210) | 98% | 0.59<br>(n=210) | 85% |
| **Mild**<br>**anemia** | 0.74<br>(n=196) | 71% | 0.52<br>(n=196) | 98% | 0.49<br>(n=196) | 94% |
| **Moderate**<br>**anemia** | 1.23<br>(n=265) | 47% | 0.52<br>(n=265) | 97% | 0.52<br>(n=265) | 91% |
| **Severe**<br>**anemia** | 1.43<br>(n=106) | 47% | 0.47<br>(n=106) | 96% | 0.43<br>(n=106) | 93% |

### Accuracy of classification of anemia using AI model, clinical examination, and Hemocue point of care test

In the test set (n=777), the sensitivity and specificity of the AI model for the presence of anemia were 70% (95%CI 66-74) and 70% (95%CI 64-76) respectively (**Table 3**). Sensitivity and specificity were higher for severe anemia vs mild, moderate or no anemia, 90% (95%CI 82-94) and 80% (95%CI 77-83) respectively.

**Table 3:** Accuracy of anemia classification by AI model, clinical examination and Hemocue venous and capillary samples in the test set (n=777)

|  | **Anemia vs. No Anemia** | **Severe/Moderate vs. Mild/Normal** | **Severe Anemia vs. Mild/Moderate/Normal** |
| --- | --- | --- | --- |
|  | Sensitivity, Specificity<br>(95% CIs) | Sensitivity, Specificity<br>(95% CIs) | Sensitivity, Specificity<br>(95% CIs) |
| **ResNet AI model**<br>(n=777) | 70 [66-74], 70 [64-76] | 74 [69-78], 72 [68-77] | 90 [82-94], 80 [77-83] |
| **Clinical examination**<br>(n=772*) | 73 [70-78], 53 [46-60] | 40 [36-45], 94 [91-96] | 31 [22-40], 100 [99-100] |
| **Hemocue (venous)**<br>(n=777) | 86 [82-88], 99 [97-100] | 79 [75-83], 99 [98-100] | 82 [74-88], 100 [-] |
| **Hemocue (capillary)**<br>(n=516)** | 90 [87-93], 83 [77-89] | 86 [81-90], 92 [89-95] | 90 [80-96], 100 [98-100] |

Note: \*5 samples in test data were labeled as “Not Sure” by the researcher. \*\*261 samples in test set did not have capillary blood samples

Clinical assessment of the presence of anemia (**Table 3**) showed sensitivity and specificity for differentiating anemia from no anemia of 73% (95%CI 70-78) and 53% (95%CI 46-60) respectively. Sensitivities for detecting severe or moderate vs mild or no anemia was lower (40%, 95%CI 36-45), but specificity was high (94%, 95%CI 91-96). Similarly, sensitivity for clinical assessment of severe vs mild, moderate or no anemia was low (31%, 95%CI 22-40) with high specificity (100%).

The performance of the model on the field-representative validation dataset remained similar to the test dataset (**Appendix 6**). One important exception was that the performance deteriorated when the images were captured in outdoor conditions where the light intensity was high (data not shown).

The accuracy of the Hemocue point of care test for classification of anemia was higher using capillary samples than venous samples (**Table 3**). Using venous samples Hemocue demonstrated sensitivity and specificity for anemia vs no anemia of 86% (95%CI 82-88) and 99% (97-100). Accuracy for differentiating severe vs mild, moderate or no anemia showed sensitivity of 82% (74-88) and specificity of 100%. The Hemocue point of care test on **capillary** samples where this was available (n=516) demonstrated sensitivity and specificity for anemia vs no anemia of 90% (95%CI 87-93) and 83% (77–89). Accuracy for differentiating severe vs mild, moderate or no anemia were higher with sensitivity of 90% (80–96) and specificity of 100% (98–100).

Receiver Operating Characteristic (ROC) curves, including areas under the curve (AUC) and operating points for the AI model, Hemocue point of care test and clinical assessment are displayed in **Figure 3**. For classifying anemia vs no anemia, the AI model showed AUC of 0.80, compared to 0.98 for Hemocue. For classifying moderate or severe vs normal Hb/mild anemia, the AUROC for the AI model and Hemocue were 0.81 and 1.0 respectively. Finally, the AUROC of the AI model and Hemocue for classifying severe anemia vs mild/moderate anemia/normal Hb, were 0.93 and 1.0 respectively.

**Figure 3:**
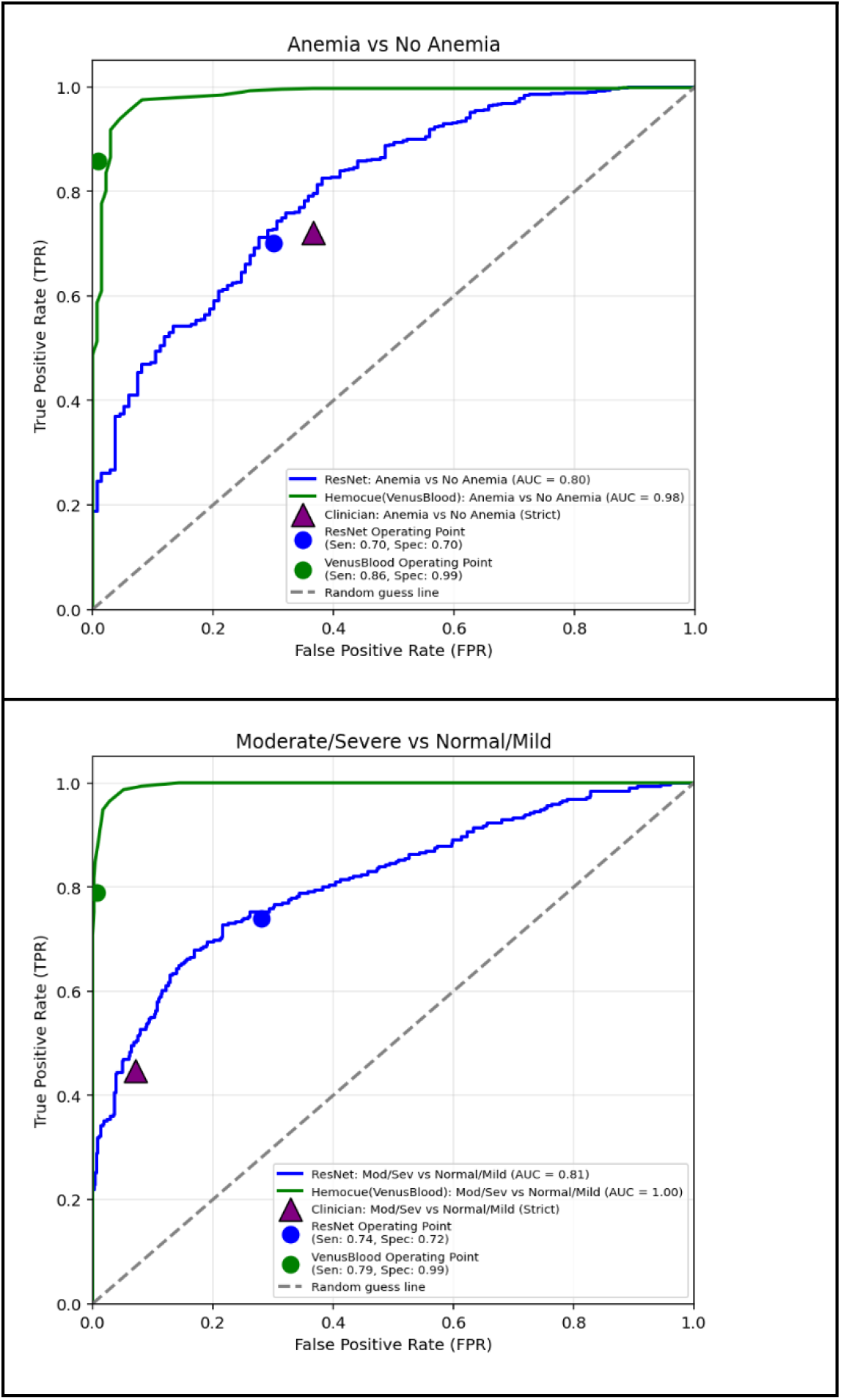
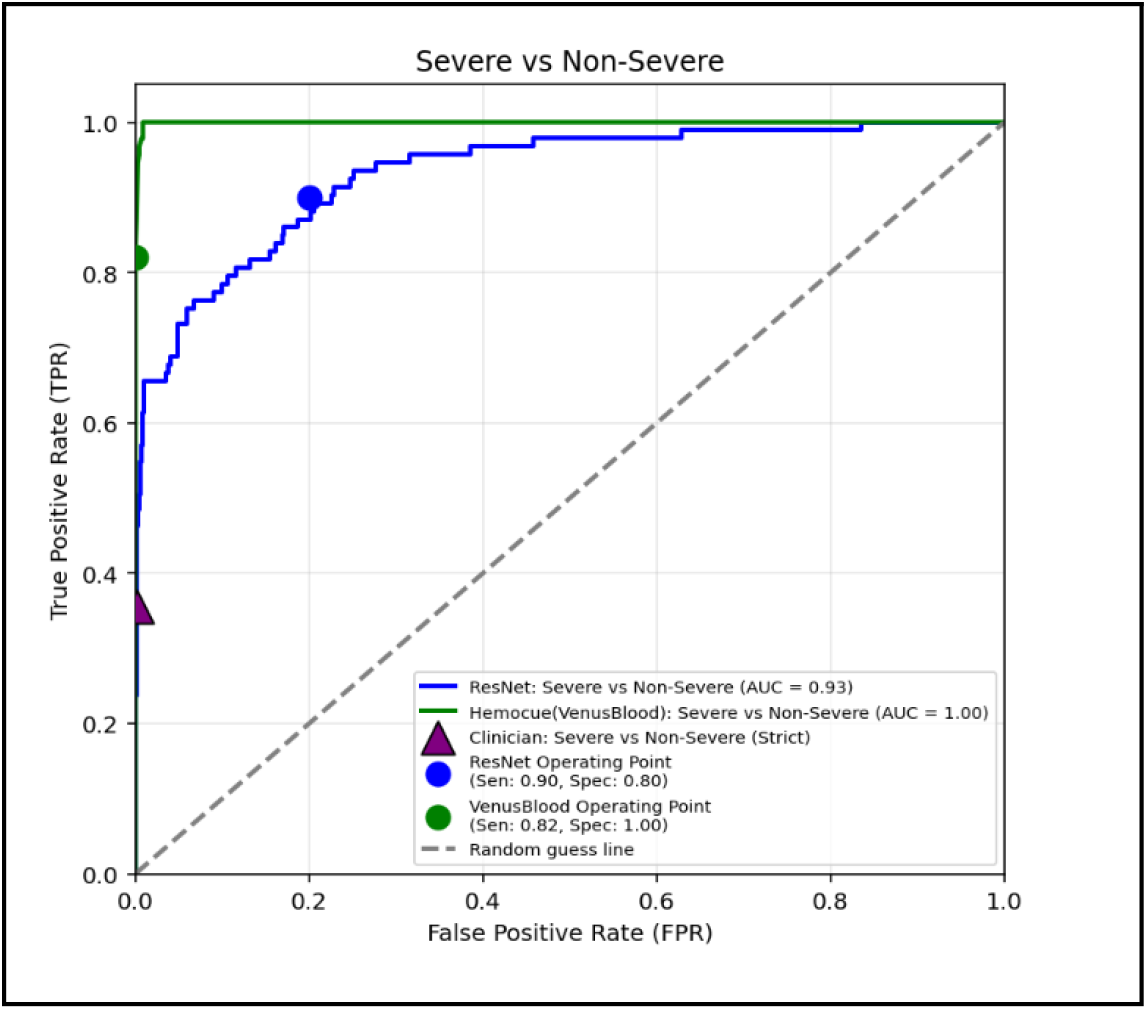
Receiver Operating Characteristic (ROC) curves and operating points for AI model, Hemocue (venous sample), and clinical examination on test set (n=777)

### Accuracy of classification of anemia using AI model across subgroups

Among subgroups, sensitivity of the AI model was higher for women than men, and specificity was higher for men than women across all three classification groups in the test set (n=777) (Table 4). Sensitivity was higher and specificity was lower among non-pregnant women for all anemia grade classifications. Similarly, for the youngest age group (1-12 yrs) sensitivity was higher and specificity lower than the older age groups for all three anemia classifications. Overall sensitivity and specificity values were similar, with overlapping 95%CI across the three classification groups (Table 4). A total of 106 (106/777, 13.6%) of participants in the test set have Sickle Cell disease, of which 42 (5.4%) had Sickle Cell Trait and 115 (14.8%) were negative, while the majority had unknown status. For those with known Sickle Cell disease present, sensitivity and specificity were somewhat higher than that of the overall sample (Table 4). Hemocue remained highly sensitive and specific across all subgroups (Appendix 7)

**Table 4:** Accuracy of anemia detection by AI model for subgroups of age, sex, pregnancy, skin tone, sickle cell status, smartphone device, light intensity in the test set (n=777)

|  |  | **Anemia vs No Anemia** | **Severe/Moderate vs Mild/Normal** | **Severe Anemia vs Mild/Moderate/Normal** |
| --- | --- | --- | --- | --- |
|  |  | Sensitivity, Specificity (95% CIs) | Sensitivity, Specificity (95% CIs) | Sensitivity, Specificity (95% CIs) |
| **Sex** |  |  |  |  |
| Female | 652 | 71 [67-75], 68 [61-74] | 76 [71-81], 69 [64-74] | 90 [82-95], 79 [75-82] |
| Male | 125 | 63 [53-73], 83 [67-94] | 63 [51-75], 90 [79-95] | 88 [67-100], 83 [76-89] |
| **Pregnancy status (among female population)** |  |  |  |  |
| Pregnant | 371 | 59 [53-65], 69 [62-77] | 63 [54-72], 74 [68-80] | 63 [39-83], 87 [83-90] |
| Not pregnant | 281 | 82 [77-86], 61 [42-76] | 84 [79-89], 54 [43-64] | 97 [91-100], 65 [58-72] |
| **Age group** |  |  |  |  |
| 0-12 years | 75 | 81 [70-89], 50 [13-86] | 84 [72-92], 47 [26-72] | 80 [0-100], 59 [46-69] |
| 0-19 years | 248 | 78 [71-83], 58 [45-73] | 77 [69-83], 58 [49-67] | 75 [53-90], 74 [68-79] |
| >19 years | 529 | 65 [61-70], 74 [67-81] | 73 [67-78], 78 [73-83] | 93 [86-97], 83 [79-86] |
| **Sickle cell disease status** |  |  |  |  |
| Disease | 106 | 84 [77-91] \| 75 [0-100] | 77 [67-86] \| 64 [36-88] | 88 [69-97] \| 54 [43-64] |
| Negative | 115 | 74 [65-83] \| 77 [57-90] | 71 [57-82] \| 67 [55-79] | 100 [-] \| 82 [73-88] |
| Not known | 514 | 66 [60-70] \| 70 [63-77] | 74 [68-79] \| 74 [69-79] | 88 [78-95] \| 84 [80-87] |
| Trait | 42 | 55 [38-72] \| 55 [25-86] | 68 [48-86] \| 70 [45-87] | 100 [-] \| 81 [65-92] |
| **Skin tone categories** |  |  |  |  |
| Very Light/Light | 1 (Light) | - | - | - |
| Intermediate | 2 | 100 [0-100] \| 100 [0-100] | 100 [0-100] \| 100 [0-100] | 0 [-] \| 50 [0-100] |
| Tan | 39 | 66 [47-82] \| 70 [31-92] | 80 [56-95] \| 95 [66-100] | 100 [-] \| 85 [71-94] |
| Brown | 592 | 69 [65-74] \| 69 [61-75] | 74 [69-79] \| 71 [66-76] | 88 [79-94] \| 79 [76-83] |
| Dark | 143 | 73 [64-81] \| 78 [63-89] | 73 [61-83] \| 71 [60-81] | 95 [75-100] \| 79 [70-85] |
| **Smartphone type** |  |  |  |  |
| SAMSUNG A05 | 276 | 63 [56-69] \| 72 [60-81] | 71 [62-78] \| 71 [64-79] | 86 [70-95] \| 80 [75-85] |
| SAMSUNG A14 | 243 | 76 [70-82] \| 68 [57-80] | 79 [70-85] \| 73 [64-80] | 93 [81-98] \| 82 [75-86] |
| SAMSUNG S24 | 258 | 71 [64-77] \| 72 [61-81] | 73 [64-80] \| 73 [66-80] | 89 [71-97] \| 77 [72-82] |
| **Light intensity** |  |  |  |  |
| Light Intensity quantile=0 (18.0 - 151.0) | 199 | 67 [59-75] \| 64 [51-77] | 77 [67-84] \| 73 [64-81] | 82 [66-94] \| 84 [78-89] |
| Light Intensity quantile=1 (151.0 - 182.0) | 216 | 72 [65-79] \| 74 [62-84] | 74 [65-82] \| 71 [63-79] | 93[74-100] \| 81 [74-86] |
| Light Intensity quantile=2 (182.0 - 211.0) | 191 | 70 [62-77] \| 66 [53-76] | 73 [61-82] \| 71 [62-79] | 100 [-] \| 78 [72-84] |
| Light Intensity quantile=3 (211.0 - 665.0) | 169 | 69 [61-76] \| 81 [63-91] | 72 [62-81] \| 74 [64-83] | 83 [63-95] \| 75 [68-82] |

### Metadata Ablation Study Results

The ablation studies which compare the performance of the baseline model (using conjunctiva and patient metadata) with single or combinations of other images like nail bed or tongue, clinical metadata, and symptoms on accuracy of Hb estimation and anemia classification are shown in **Appendix 8**. No single or combination of image modalities provided lower MAE than the baseline model. A model that combined conjunctiva with patient metadata and child or adult symptoms did provide a lower MAE compared to the baseline model, of 1.099 mg/dL vs 1.124 mg/dL, AUC for anemia vs no anemia of 0.937 vs. 0.829, moderate or severe anemia vs normal Hb or mild anemia of 0.839 vs. 0.828, and severe anemia vs not severe anemia of 0.973 vs 0.968, respectively.

## Discussion

### Main findings

The AI model’s estimation of Hb value in the final test set demonstrated an overall MAE of 1.12 g/dL, while the sensitivity and specificity of the AI model for the presence of anemia were 70% (95%CI 66-74) and 70% (95%CI 64-76) respectively. Sensitivity and specificity were higher for classifying severe anemia vs mild, moderate or no anemia, 90% (95%CI 82-94) and 80% (95%CI 77-83) respectively, which is important given that individuals with severe anemia are most likely to benefit from point of care screening, and time sensitive interventions to facilitate prompt treatment(3, 5). We found that a single image of the conjunctiva along with a person’s age, sex and pregnancy status provided marginally lower accuracy than a more complex model involving these features as well as the presence or absence of symptoms in adults (9 symptoms) and in children (4 symptoms). The marginal gain of accuracy with the more complex model, requiring in any future use for this additional symptom data to be accurately reported and entered, was not considered a worthwhile tradeoff, and so the main results focused on the simpler model without these symptom inputs. While we explored using different image types in addition to, or instead of the conjunctiva, as well as PPG signals, these yielded comparable or lower accuracy, leading to the conclusion to use conjunctival images alone as the optimal body site images in this study, for anemia detection.

Accuracy results did not vary significantly across a range of skin tones, and smartphone models, which implies that findings may be generalisable to other populations, and not limited to users of high-end smartphones. The AI model performance was sustained on a field-representative validation dataset generally; though high intensity of illumination in outdoor settings lowered the accuracy. The model struggled most with the boundary between normal Hb and mild anemia. This may be due to the subtle chromatic differences in the conjunctiva between mild anemia and no anemia that are smaller than the variance caused by a smartphone’s auto-white-balance settings. Even with high-end mobiles like the Samsung S24, the “auto-white balance” of smartphone cameras can shift the perceived redness of the conjunctiva. Our use of a “white paper reference” during capture was essential for normalizing these color shifts. Finally, in children (ages 1–5 years), we noted that data rejection difficulties arose primarily due to motion artifacts in capturing a clear, retracted eyelid image, suggesting potential operational challenges in capture and use of conjunctiva for this age group.

The AI model’s performance in anemia classification was compared to that of the clinician examining the patient, who was not blinded to their Hb results. We noted overall lower clinician sensitivity particularly for the severe or moderate vs mild or normal Hb categories, which limits the value of clinical examination for screening. However we noted that specificity of the clinical examination was higher than that of the AI model. Not surprisingly, the Hemocue point of care device provided highly accurate classification, particularly with the capillary samples, with higher sensitivity and specificity than the AI model across all categories. The fact that the AI model using non-invasive smartphone images had substantially higher sensitivity for severe anemia than clinician assessment of physical signs, suggests potential for augmenting screening using this accessible, objective approach of Hb estimation and anemia classification.

### Comparison with literature

Many studies have used machine learning with smartphone images of various body sites, in attempts to estimate Hb levels or classify anemia. The challenge with contextualizing within this body of research is that studies are conducted in highly variable populations in terms of size, demographics, distribution of Hb values, as well as varying proportions with ground truth Hb measurement, varying levels of validation, and different technical approaches (33). Comparing findings is further limited by highly variable reporting of many of these studies.

A large study of AI-augmented app using fingernail images, estimated Hb with MAE of +/-0.72 g/dL testing 200,000 U.S. users, and reported a sensitivity of 89% and specificity of 93% at an anemia cutoff of 12.5 g/dL, with MAE improving from 1.36 to 0.74 g/dL after adding in user details, though with relatively few participants with moderate or severe anemia (34). Lip mucosa has been examined in small cohorts, showing variable classification accuracies, for example one study of 138 individuals found overall accuracy ranging from 75 to 96% depending on the model used (35). Lips and the oral mucosa can sometimes be colored by substances such as Betel nut or tobacco, which could limit generalisability to some communities.

Conjunctival imaging, the modality most directly relevant to the present study, has produced a heterogeneous body of evidence. A convenience sample of 435 patients in the emergency department estimated Accuracy of Hb estimation was 75.4% (95% CI 71.3-79.4) for all categories of anemia (36), A radiomics-based approach applied to over 12,000 grayscale conjunctival images from 565 children used machine learning without deep learning (37). Notably, the NiADA smartphone application, validated across 556 pediatric, pregnant, and adult participants in India, demonstrated inconsistent performance across subgroups — sensitivity/specificity of 75.8%/53.8% in adult females versus 23.8%/90% in pregnant women — underscoring the challenge of generalizing conjunctiva-based models across physiologically distinct populations (38, 39).

Collectively, this literature suggests that while image-based hemoglobin estimation is technically feasible across multiple body sites, model performance remains highly dependent on population characteristics, image standardization, and validation methodology.

### Strengths and limitations

The study had several strengths. First, the accuracy of the final models to differentiate severe anemia from moderate or mild was within the target product profile suggested by expert groups consulted before the initiation of the study. Second, the dataset collected for this study provides the largest dataset available that covers a range of values of Hb, surpassing existing alternatives by offering three image modalities (conjunctiva, fingernails, tongue), clinically audited metadata, and a broad demographic range from children to adults. This dataset has a large number of cases at the extremes of Hb values which is unique. Third, we recruited participants from populations in India which provides real world data on various skin tones from a mainly rural population. Fourth, we examined the use of three different smartphones, to provide evidence of the impact of device cost/sophistication on performance of algorithms. This is important given resource constraints for deploying higher end smartphones in healthcare settings in lower resource areas. Lastly, this is to our knowledge the only study that explored the multi-modal approach using images of tongue, conjunctiva and nailbeds.

However, we note several limitations. First, while the accuracy of the final model for classification of anemia was higher than other studies using this technology, they remain lower than the invasive point of care Hb devices which are used in many lower resource settings, including that of the Hemocue test as noted in our study. Specifically, Hb levels were over-predicted for severe class and under-predicted for normal/mild/moderate classes. Second, the population studied here, while drawn from the rural population of this region who were attending the hospital, may not be representative of individuals with different skin tones, or factors that might affect images of the conjunctiva, tongue or nailbeds (e.g. skin markings, eye irritation). Finally, the research team collecting data on participants were not blinded to Hb results, therefore their assessment of presence of symptoms, signs and classification of anemia is potentially subject to bias. This may have led to higher accuracy for the clinical assessment of anemia presence, although overall the accuracy of clinical assessment was in line with published studies (7, 8). Finally, our sampling method was non-consecutive sampling, which is known to overestimate estimates of accuracy in studies of diagnostics, but allowed us to collect data on a range of Hb levels which is essential for developing and testing models (40).

### Implications for researchers, policy makers, others

Our findings have several implications for clinicians and policy makers. Based on the study results, it may be viable to consider deploying a smartphone based severe anemia detection tool as part of routine screening in populations similar to those studied here. Two use cases could be considered: first, screening patients who decline invasive blood testing or invasive blood testing using a noninvasive alternative. Second, a cascaded approach using a smartphone tool as first level screening, followed by use of point of care blood test to confirm suspected anemia cases. We believe this cascaded approach would likely minimize the cost of large scale screening without missing severe anemia cases. Additional modelling and understanding of human behavior is needed to determine the most appropriate step-wise use of this approach.

Further research is necessary using larger datasets from diverse populations. Additional technology efforts may be needed to provide user interfaces suitable for frontline healthcare workers to ensure capture of good quality images of conjunctiva as well as developing on-device models for remote territories that lack internet connection. Although the model is performing well for screening of severe anemia, further training of the model is necessary to improve the accuracy of the model for other severity classes of anemia. Clinical validation of the model in real life community settings is required to assess accuracy in deployment settings. Beyond addressing these limitations, several translational imperatives must be met. Most non-invasive optical platforms remain investigational Software as a Medical Device (SaMD), whereas FDA clearance currently remains restricted to minimally invasive capillary tests (e.g., AnemoCheck Home). Future clinical adoption requires moving beyond binary classification to continuous quantitative Hb regression (g/dL), external validation in pregnant and multi-ethnic cohorts, and hardware-agnostic sensor calibration. Overcoming these translational barriers is essential to accurately evaluating the cost-utility, clinical effectiveness, and practical feasibility of deploying smartphone based models in real world settings. To conclude, we studied the use of noninvasive smartphone based imaging of various body sites for anemia classification and found the conjunctiva alongside simple demographic information provided acceptable accuracy. Further studies and validations will be needed to improve performance and examine how best to integrate such tools into real screening workflows

## Data sharing statement

The dataset used in this study is publicly available on Hugging Face^25^. The code developed for this study, including the scripts used for model development and evaluation, is publicly available at https://github.com/Google-Health/google-health/tree/master/non-invasive-anemia-ai to facilitate reproducibility.

## Acknowledgments

We thank all the patients who participated in this study. We would like to thank the National Institute of Nutrition-ICMR) for technical guidance, including reference standard validation. We would like to acknowledge Argusoft India Limited for the development of the data collection mobile application.

