## Supplementary Appendix for "Smartphone-based Non-Invasive Classification of Anemia Severity using Deep Learning: A Cross-Sectional Study in Rural India"

### Appendix 1: Method of capturing image of conjunctiva and nailbed

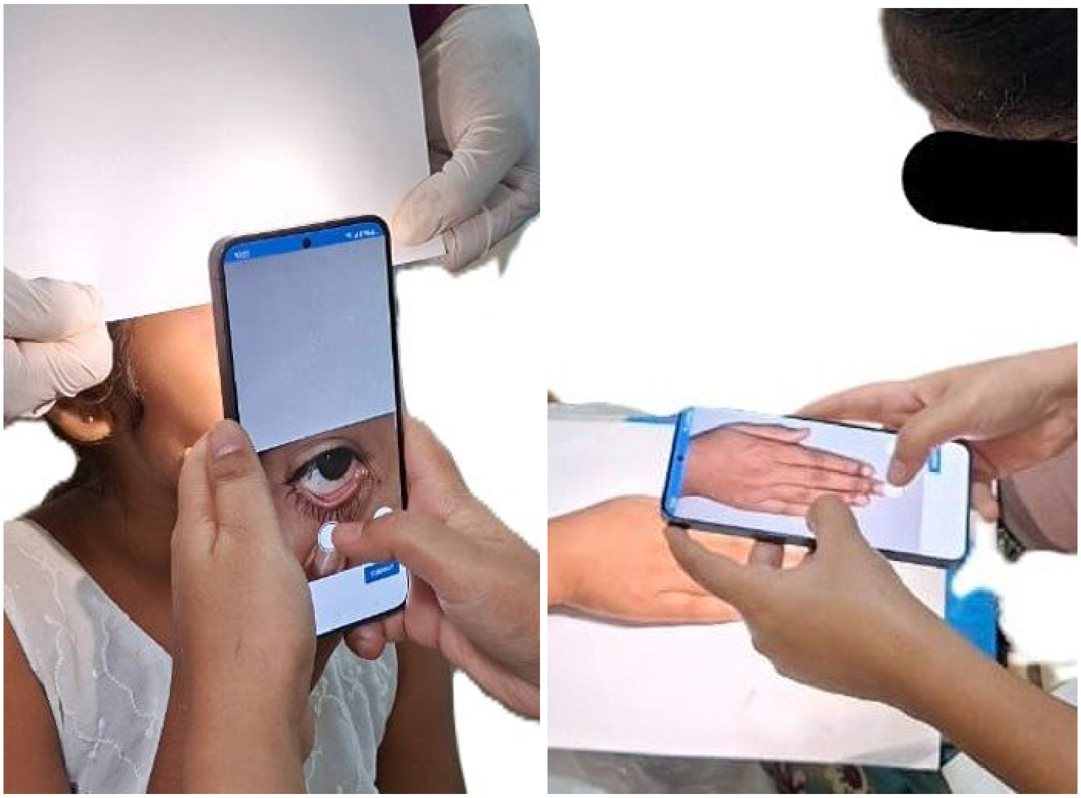

### Appendix 2 Overall Model Architecture

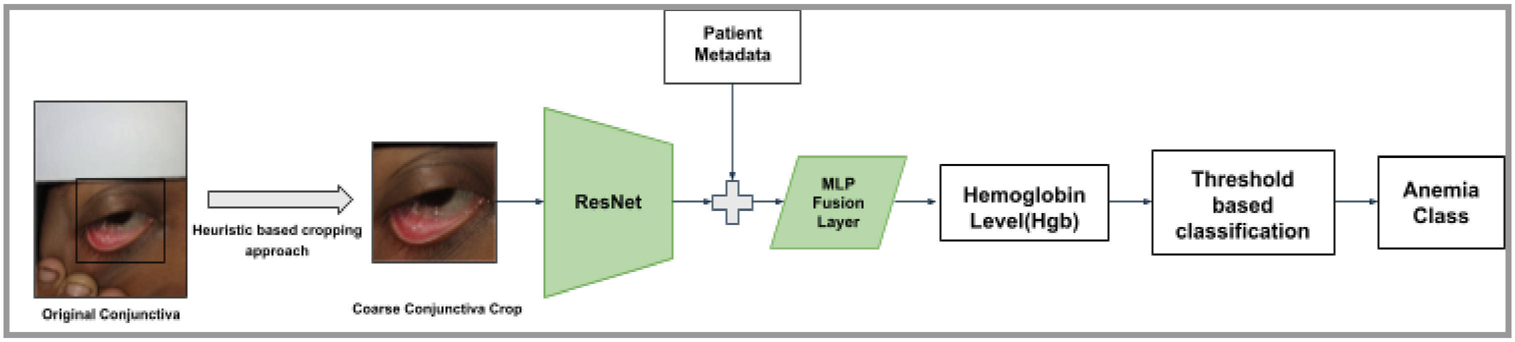

### Appendix 3  A Two Stage ResNet Block

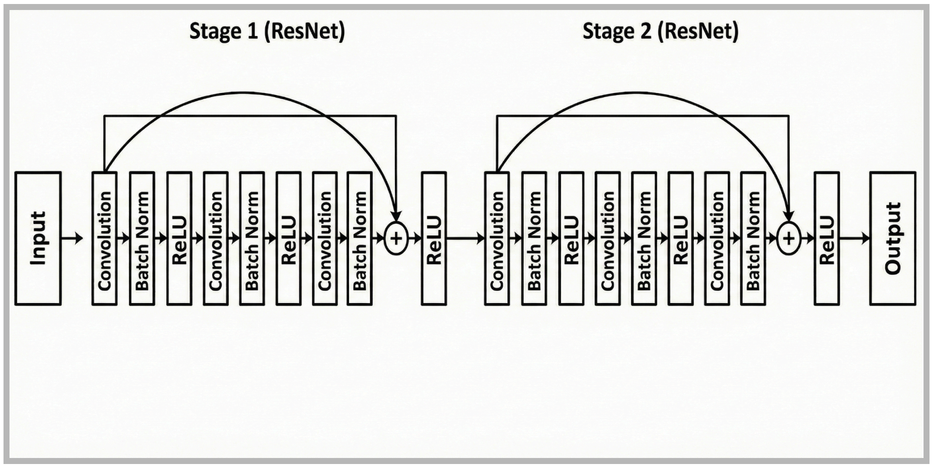

**Appendix 4: Hemoglobin cutoffs to define anemia severity** (29)

| Population | No Anemia (g/dL) | Mild Anemia (g/dL) | Moderate Anemia (g/dL) | Severe Anemia (g/dL) |
|---|---|---|---|---|
| Children 6–23 months of age | ≥ 10.5 | 9.5–10.4 | 7.0–9.4 | < 7.0 |
| Children 24–59 months of age | ≥ 11.0 | 10.0–10.9 | 7.0–9.9 | < 7.0 |
| Children 5–11 years of age | ≥ 11.5 | 11.0–11.4 | 8.0–10.9 | < 8.0 |
| Children 12–14 years of age | ≥ 12.0 | 11.0–11.9 | 8.0–10.9 | < 8.0 |
| Adults, 15-65 years, Non-pregnant women | ≥ 12.0 | 11.0–11.9 | 8.0–10.9 | < 8.0 |
| Adults, 15-65 years, men | ≥ 13.0 | 11.0–12.9 | 8.0–10.9 | < 8.0 |
| Pregnant women (1st trimester) | ≥ 11.0 | 10.0–10.9 | 7.0–9.9 | < 7.0 |
| Pregnant women 2nd trimester) | ≥ 10.5 | 9.5–10.4 | 7.0–9.4 | < 7.0 |
| Pregnant women (3rd trimester) | ≥ 11.0 | 10.0–10.9 | 7.0–9.9 | < 7.0 |

#### Appendix 5: Model Configuration

| Category | Component / Parameter | Configuration Details |
|---|---|---|
| **Data Specifications** | **Model Inputs** | Conjunctiva Images + Patient Metadata (age, sex, pregnancy status) |
| **Architecture** | **Backbone Structure** | ResNet (2 Stages: [1, 1] Blocks) |
| | **Task Type** | Regression (Hb prediction) |
| **Optimization** | **Optimizer** | Adam |
| | **Learning Rate Scheme** | 0.013 Initial LR (with Exponential Decay) |
| | **Loss Function** | Huber Loss (Robust to outliers) |
| **Execution** | **Training Duration** | 150 Epochs |
| | **Batch Size** | 32 Samples |
| **Data Strategy** | **Imbalance Handling** | Inverse Bin Frequency Reweighting (Applied to Hgb Target) |

#### Appendix 6: Accuracy on data collected from field-representative collection set

| | Sensitivity (95%CI) | Specificity (95%CI) | Severe anemia cases missed (%) |
|---|---|---|---|
| **Anemia** (n= 319) **vs** **No Anemia** (n=197) | 66 [61-71] | 69 [62-75] | 4.5 |
| **Severe/Moderate** (n=188) **vs** **Mild/Normal** (n=328) | 73 [67-80] | 69 [64-74] | 4.5 |
| **Severe Anemia** (n=44) **vs** **Mild/Moderate/Normal** (n=472) | 95 [84-100] | 78 [74-82] | 4.5 |

*Note: Performance metrics were evaluated across the total test pool (N = 516 images) obtained from n = 429 unique participants. 3 images from 29 participants each is taken at different light intensities and physical location)*

**Appendix 7. Accuracy of Hemocue (venous samples) for anemia detection for overall population, and subgroups of age, sex, pregnancy in the test set (n=777)**

| | **Anemia vs No Anemia** | **Severe/Moderate vs Mild/Normal** | **Severe Anemia vs Mild/Moderate/Normal** |
|---|---|---|---|
| | Sensitivity, Specificity (95%CIs) | Sensitivity, Specificity (95%CIs) | Sensitivity, Specificity (95%CIs) |
| **Overall population (n=777)** | | | |
| | 86 [82-88], 99 [97-100] | 79 [75-83], 99 [98-100] | 82 [74-88], 100 [-] |
| **Sex** | | | |
| **Female (n=652)** | 85 [81-88], 99 [97-100] | 77 [73-82], 99 [98-100] | 79 [69-86], 100 [-] |
| **Male (n=125)** | 90 [82-95], 97 [87-100] | 88 [77-95], 99 [93-100] | 100 [-], 100 [-] |
| **Pregnancy status (among female population)** | | | |
| **Pregnant (n=371)** | 78 [72-82], 99 [96-100] | 67 [58-75], 100 [98-100] | 53 [29-75] , 100 [-] |
| **Not pregnant (n=281)** | 92 [88-95], 100 [-] | 84 [78-88], 99 [93-100] | 86 [76-93], 100 [-] |
| **Age group** | | | |
| **0-12 years (n=75)** | 90 [79-95], 100 [-] | 88 [76-95], 100 [-] | 80 [0-100], 100 [-] |
| **0-19 years (n=248)** | 87 [82-91], 100 [-] | 78 [71-85], 99 [95-100] | 85 [63-100], 100 [-] |
| **>19 years (n=529)** | 85 [81-88], 99 [96-100] | 79 [74-85], 99 [98-100] | 81 [73-89], 100 [-] |

**Appendix 8: Incremental Multimodal ablation study on validation dataset (n=776)**

| | Feature | ML Model | MAE (g/dL) | Anemia vs. No Anemia AUC | Moderate or Severe anemia vs. Normal Hb or Mild Anemia AUC | Severe anemia Vs Not severe Anemia AUC |
|---|---|---|---|---|---|---|
| **Baseline*** | conjunctiva + patient metadata | Resnet | 1.124 | 0.829 | 0.828 | 0.968 |
| **Single Image Modality** | conjunctiva | Resnet | 1.221 | 0.819 | 0.831 | 0.967 |
| | tongue | Resnet | 1.824 | 0.656 | 0.665 | 0.810 |
| | Nails open | Resnet | 1.504 | 0.749 | 0.761 | 0.904 |
| | Nails closed | Resnet | 1.575 | 0.732 | 0.745 | 0.875 |
| **Two Image Modality** | conjunctiva + tongue | Resnet | 1.222 | 0.812 | 0.821 | 0.966 |
| | conjunctiva + nails open | Resnet | 1.144 | 0.832 | 0.844 | 0.966 |
| | conjunctiva + nails closed | Resnet | 1.202 | 0.804 | 0.831 | 0.978 |
| | tongue + nails open | Resnet | 1.484 | 0.760 | 0.761 | 0.901 |
| | tongue + nails closed | Resnet | 1.529 | 0.724 | 0.745 | 0.881 |
| | Nails open + nails closed | Resnet | 1.446 | 0.755 | 0.767 | 0.907 |
| **Three & All Image Modality** | conjunctiva + tongue + nails open | Resnet | 1.136 | 0.825 | 0.840 | 0.978 |
| | conjunctiva + tongue + nails closed | Resnet | 1.173 | 0.804 | 0.823 | 0.967 |
| | conjunctiva + nails | Resnet | 1.159 | 0.813 | 0.833 | 0.971 |

| | | | | | | |
|---|---|---|---|---|---|---|
| | open + nails closed | | | | | |
| | tongue + nails open + nails closed | Resnet | 1.389 | 0.765 | 0.782 | 0.913 |
| | all image modalities | Resnet | 1.139 | 0.822 | 0.836 | 0.968 |
| **Best Image Modality + Metadata*** | conjunctiva + patient metadata + child symptoms + adult symptoms | Resnet | 1.099 | 0.837 | 0.839 | 0.973 |
| **Only Metadata Experiments** | Age + Gender + Pregnancy status | ElasticNet | 1.634 | 0.571 | 0.611 | 0.719 |
| | Age+ Gender + Pregnancy status | Random Forest | 1.610 | 0.618 | 0.625 | 0.737 |
| | child symptoms + adult symptoms | ElasticNet | 1.631 | 0.564 | 0.608 | 0.748 |
| | child symptoms + adult symptoms | Random Forest | 1.616 | 0.564 | 0.608 | 0.768 |
| **Test Dataset(n=777)**** *Performance of the two selected combinations(*) on the held-out test dataset* | | | | | | |
| **Baseline** | conjunctiva + patient_metadata | Resnet | 1.111 | 0.798 | 0.824 | 0.927 |
| **Best Image Modality + Metadata** | conjunctiva + patient_metadata + child symptoms + adult symptoms | Resnet | 1.118 | 0.810 | 0.819 | 0.934 |

***Adult symptoms:*** *lack of energy, shortness of breath, fast heart beat, losing weight, chest pain, dizziness, headache, leg cramps, brittle nails or hair.*

***Child symptoms:*** *looks pale, losing weight, tiredness, breathing difficulty.*

***Patient Metadata:*** *Age, gender, pregnancy status*

****Note:** conjunctiva + patient_metadata was selected as the baseline due to its smaller input set. Other combination extend it with child/adult symptoms. Test-set performance is reported for both these combinations to compare their generalization performance.
